# Prescription of nutritional interventions to HIV patients in Dar es Salaam, Tanzania

**DOI:** 10.1101/2023.11.12.23298397

**Authors:** Ajibola I Abioye, Hellen Siril, Aisa Mhalu, Nzovu Ulenga, Wafaie W Fawzi

## Abstract

**Background:** Anemia and micronutrient deficiencies are common among people living with HIV (PLHIV). There are no current guidelines from the World Health Organization (WHO) regarding whether supplements are recommended or not. We sought to assess the practices with respect to nutritional supplementation among clinicians providing care to people living with HIV in Dar es Salaam, Tanzania.

**Methods:** We conducted a cross-sectional survey at clinics providing care to PLHIV in Dar es Salaam, Tanzania. All healthcare workers with prescribing responsibility were invited. Self-administered questionnaires were used to collect information about participants’ demographic and professional characteristics, and their approach to making decisions regarding the prescription of nutritional interventions. Descriptive analyses regarding prescribing patterns and associated factors were done.

**Results:** Two hundred and fifty-four clinicians participated in the survey. They were clinical officers (65%), medical assistants (21%) or medical doctors (13%), and attended to 30 patients (IQR: 10, 100) on average, per week. While the majority usually prescribed iron and multivitamin supplements (79% and 76%, respectively), only 33% usually prescribed ready-to-use therapeutic foods (RUTF). The decision to prescribe nutritional supplements were typically guided by patients’ clinical condition and laboratory test results. Pallor was the most commonly considered clinical feature across patient subgroups. Most participants commenced supplementation when hemoglobin concentration was ≤10g/dl. Clinicians who attended to between 10 and <100 patients or believed in the need for universal iron supplementation for pregnant PLHIV were more likely to prescribe iron supplements compared to counterparts who attended to <10 patients weekly or who did not believe in the need for universal iron supplementation for pregnant PLHIV respectively.

**Conclusion:** Clinicians frequently prescribe nutritional supplements, with considerable variation in how they decide whether and how to.

## Introduction

Mortality among people living with HIV (PLHIV) has declined considerably in the last 15 years, due to wider access to highly active antiretroviral therapy (HAART) regimens^(1; 2)^. Despite this tremendous progress, 7 – 13% of HIV-infected patients in sub-Saharan African countries still die in the first year of HAART^(3; 4; 5)^. Evidence among pre-HAART and HAART era suggests that poor nutrition is likely to be partly responsible for the slow decline in mortality rates. For instance, anemia is very common among PLHIV, with a prevalence of 85% in some settings, and iron deficiency is responsible for a considerable proportion of cases^(6; 7)^. Anemia is associated with a greater risk of all-cause mortality, disease progression, and tuberculosis treatment failure^(8; 9; 10)^. Micronutrient deficiencies are also common. Eighty-six percent of participants in one cross-country study had at least one micronutrient deficiency and 57% had deficiencies of two or more micronutrients^(11)^. Micronutrient deficiencies are also associated with higher risk of HIV disease progression, treatment failure, and mortality^(12; 13; 14)^. Micronutrients reduce oxidative stress and improve the production of immune cells and antibodies, thereby protecting the integrity of the oral and gastrointestinal mucosa, enhancing systemic immune function, and reducing viral replication^(13; 15; 16)^.

The current guidelines from the World Health Organization (WHO) neither recommends nor discourages the use of multivitamin or iron supplements^(17)^. This is partly due to the significant equipoise regarding the safety and effectiveness of some of these interventions. Iron supplementation is well-known to prevent and treat anemia in the general population, and is recommended to pregnant women including HIV-infected pregnant women^(17)^. Iron supplementation is however potentially harmful in the context of infections and a recent large cohort study in Tanzania reported that the risk of mortality among patients who received iron supplementation at initiation of ART may be as high as 3.8 to 6.3-fold^(18)^. This risk arises because elevated iron status, determined by serum ferritin measurement, has been shown to be associated with a 2 – 3 fold greater risk of all-cause mortality^(19; 20)^. RCTs^(21; 22; 23; 24)^ that have examined the impact of iron supplement use have tended to have small sample sizes, but have reported significant reduction in anemia and iron deficiency^(23)^, improvement in immunologic status^(23)^, but no effect on viral load^(22)^. In the pre-ART era, large randomized trials demonstrated that multivitamin supplementation improved the risk of hematologic outcomes, disease progression and death^(13; 25)^, and a recent prospective cohort study among PLHIV on HAART also found that multivitamin supplementation may be protective^(26)^.

Anecdotal information suggests that clinicians continue to prescribe these nutritional interventions, but poor supply chain management and stock-outs have been important barriers to access ^(27)^. Understanding the extent of use of iron and multivitamin supplements may guide the design of large well-powered studies to re-examine the safety and effectiveness of these interventions in the context of HAART. We conducted a cross-sectional study to investigate the prescription patterns of micronutrient supplements among clinicians providing care to people living with HIV in Dar es Salaam, Tanzania. The specific objectives were to describe the reasons that clinicians providing care to PLHIV commence and discontinue nutritional interventions, to clarify the role of biomarker testing to guide the commencement and discontinuation of nutritional interventions at HIV Care and Treatment clinics in Dar es Salaam, Tanzania, and to identify barriers to the provision of nutritional interventions at HIV Care and Treatment clinics in Dar es Salaam, Tanzania

## Methods

### Study location and participants

The study was conducted at HIV care and treatment clinics (CTC) and Prevention of Mother to Child Transmission of HIV (PMTCT) clinics in Dar es Salaam between November and December 2020. The Management and Development for Health (MDH), Dar es Salaam, Tanzania provides operational support to these clinics to facilitate the care of PLHIV in Dar es Salaam. Participants in the study were health workers with prescribing responsibility at these clinics. Therefore, physicians and medical assistants will be invited to participate. There are approximately 200 clinics providing care to PLHIV in Dar es Salaam.

### Study design

This was a cross-sectional study among clinicians and medical assistants who directly provide care to PLHIV at the selected MDH-supported clinics across Dar es Salaam. All eligible clinicians would be invited to participate.

Given that there are no prior studies that have examined prescribing patterns for nutritional supplementation among PLHIV, we assumed a prevalence of 50% and a precision of 5%, in a population of approximately 300 eligible clinicians. We determined that an estimated sample size of 152 will suffice. The desired sample size for the study was therefore 183, to allow for 20% non-response.

### Study procedures

Study personnel approached physicians with informed consent forms written in Kiswahili, the language Tanzanians most commonly converse in, with copies in English for those who may prefer it. The consent forms provided detailed explanation of the purpose of the study, the approach to data collection, and their freedom to participate or otherwise. Following consenting, data was collected using netbooks or smartphones, on which an anonymous Qualtrics web survey form had been loaded. The study supervisor used a list of all physicians with active employment at these clinics to track the distribution and retrieval of paper copies of informed consent forms and study questionnaires. No identifiable data were collected on the questionnaires, and there was no record linking the individual physician to the study questionnaire.

### Study Instrument

The study questionnaire was in 4 sections. Section 1 had questions about participants’ demographic and professional characteristics. This section collected information on participants’ district, facility type, age, professional qualification (clinical officer, medical assistant or medical doctor), time since graduation (years), and a self-reported estimate of the number of patients attended to in the previous week. Sections 2 – 4 had similar questions, repeated for iron supplements, multivitamin supplements and ready-to-use therapeutic foods (RUTF). These sections had questions about whether nutritional interventions were usually prescribed, the frequency of prescription in the past week, the usual dose of prescription, and questions exploring the reasons for prescribing the nutritional interventions. Additional questions asked about hemoglobin testing practices to initiate or discontinue nutritional interventions, as well as barriers to prescribing the supplements.

### Statistical Analysis

The distribution of participants’ demographic and professional characteristics were described using frequency and percentages for categorical variables, and medians (with interquartile range (IQR)) for continuous variables. The responses to questions in sections about prescription practices were described using frequency and percentages, for each of iron supplements, multivitamin supplements and ready-to-use therapeutic foods (RUTF). Univariate log-binomial regression models estimated the probability that clinicians self-report prescribing nutritional interventions in relation to demographic and professional characteristics. Models were not adjusted for any covariates.

93% of participants had complete data on the demographic and professional characteristics, but there was substantial item missingness. No imputation was done for item missingness, and the number of respondents are, instead, reported for all items.

### Institutional ethical clearance

Institutional ethical clearance was obtained from the Tanzania National Institute of Medical Research (NIMR) and the Harvard T.H. Chan School of Public Health’s (HSPH) Institutional Review Board. The Tanzania National Institute of Medical Research (NIMR) regulates the conduct of medical and public health research in Tanzania.

## Results

Two hundred and fifty-four clinicians participated in our survey **(Table 1)**. Most were clinical officers (65%), and the others were medical assistants (21%) or medical doctors (13%). They worked at facilities at all five districts in Dar es Salaam: Kinondoni (31%), Ilala (26%), Temeke (26%), Ubungo (13%) and Kigamboni (3%); and there were mostly at care and treatment (CTC) facilities (64%). The median (IQR) age of the participants was 43 (34, 49), and most were >40 y (62%). The time since they graduated was 13 years (IQR: 5, 21), on average, and they had been working at the MDH for 5 years (IQR: 2, 11), on average. The clinicians attended to 30 patients (IQR: 10, 100) on average, per week.

**Table 1.** Participant characteristics, N=254.

| Characteristic | N (%) |
| --- | --- |
| District |  |
| Ilala | 66 (26%) |
| Kigamboni | 7 (3%) |
| Kinondoni | 80 (31%) |
| Temeke | 67 (26%) |
| Ubungo | 34 (13%) |
| Facility type |  |
| ANC | 4 (2%) |
| CTC | 162 (64%) |
| TB/HIV clinic | 21 (8%) |
| Other | 67 (26%) |
| Age |  |
| Median (IQR) | 43 (34, 49) |
| <30 | 32 (13%) |
| 30 – <40 | 59 (25%) |
| 40 - <50 | 54 (23%) |
| >50 | 94 (39%) |
| Qualification |  |
| Clinical officer | 166 (65%) |
| Medical assistant | 54 (21%) |
| Medical doctor | 34 (13%) |
| Number of years since graduation |  |
| Median (IQR) | 13 (5, 21) |
| 1 – 10 | 111 (43%) |
| 11 – 20 | 67 (26%) |
| 21 – 30 | 58 (23%) |
| 31 – 40 | 13 (5%) |
| 41 – 50 | 4 (2%) |
| Years since starting working with PLHIV |  |
| Median (IQR) | 5 (2, 11) |
| 0 – 5 | 132 (52%) |
| 6 – 10 | 56 (22%) |
| 11 – 16 | 66 (26%) |
| Number of patients seen previous week |  |
| Median (IQR) | 30 (10, 100) |
| <10 | 44 (18%) |
| 10 - <30 | 76 (31%) |
| 30 - <60 | 34 (14%) |
| 60 - <100 | 27 (11%) |
| >100 | 67 (27%) |

Most clinicians reported that they usually prescribed iron supplements (N=201; 79%) and multivitamin supplements (N=193; 76%). Only 33% (N=87) usually prescribe RUTF, while 13% (N=33) reported they usually counselled their patients regarding their nutrition **(Figure 1)**. When asked to estimate the proportion of patients they had prescribed supplements to in the past week, clinicians reported a median (IQR) of 17% (6%, 50%) for iron supplements, 25% (IQR: 5%, 70%) for MMS, and 0% (0%, 1%) for RUTF.

**Figure 1.**
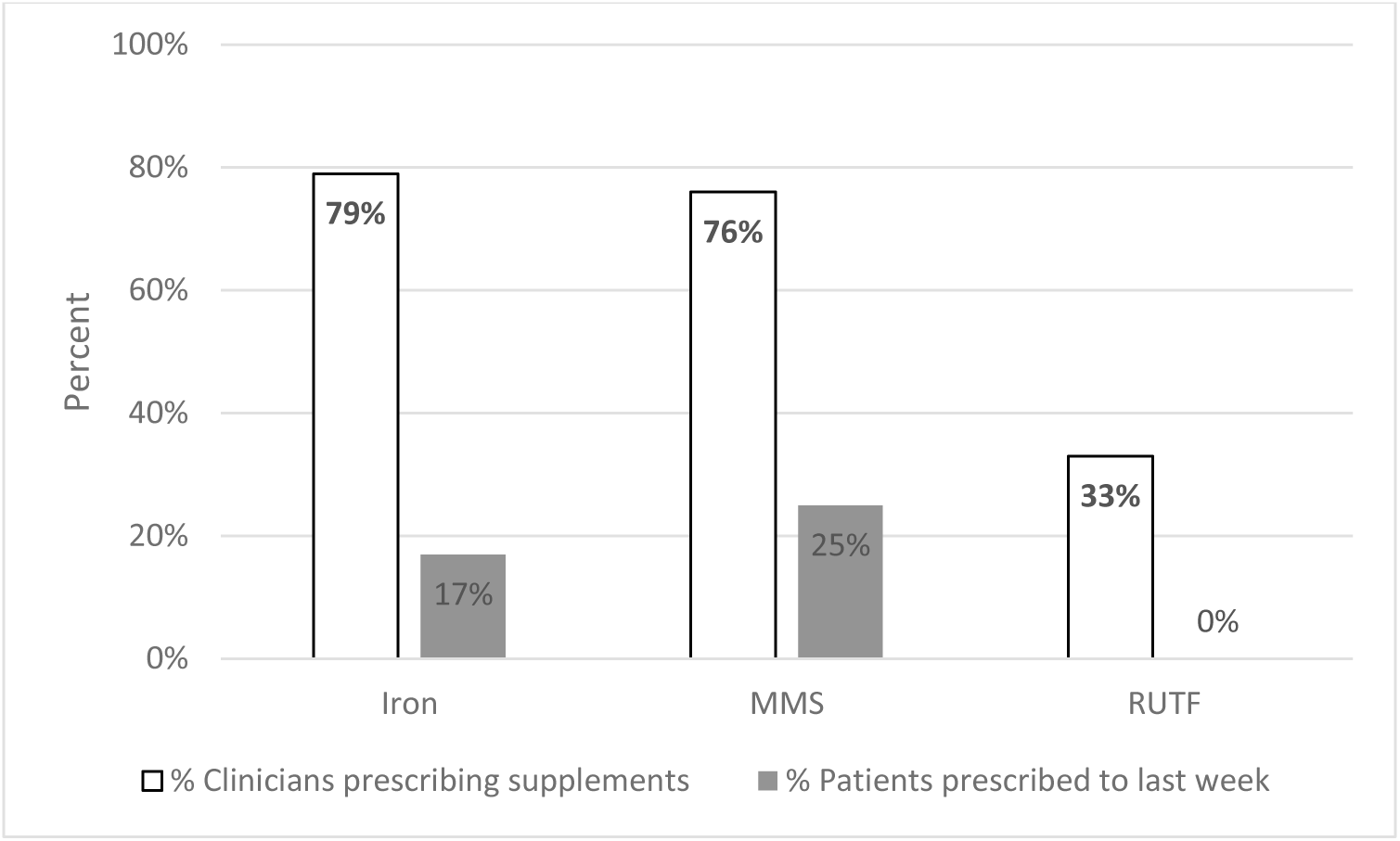
Proportion of clinicians who usually prescribed nutritional supplements.

The 201 clinicians who reported usual iron supplement prescription **(Table 2)** usually prescribed iron supplements once daily (69%) for a median of 30 days (IQR: 28, 30). Patients’ clinical condition (53%) and laboratory test results (46%) were the primary reasons for prescribing the supplements. Pallor was the most commonly considered clinical feature among non-pregnant adults (84%), children (87%) and individuals co-infected with TB (82%), though other clinical signs and symptoms were considered too. Only 82% of the clinicians believed that all pregnant PLHIV should receive iron supplements. The overwhelming majority of clinicians (98%) considered hemoglobin tests when deciding whether to prescribe, though they used a wide range of hemoglobin cutoffs to initiate iron supplements: <11 g/dl (14%), <10 g/dl (27%), <8.5 g/dl (36%) or <7 g/dl (20%). They would recommend blood transfusion when hemoglobin was lower than 7 g/dl (20%) or 8.5 g/dl (13%), or using other cutoffs in the presence of suggestive clinical symptoms or signs (29%). In addition, 85% considered higher hemoglobin cutoffs above which iron supplements should be discontinued.

**Table 2.** Prescribing pattern for iron supplements (N=201)

| Variable | n (%) |
| --- | --- |
| Number of patients prescribed iron supplements in the last week |  |
| More than 20 | 22 (11%) |
| 10 to 20 | 32 (16%) |
| 5 to 9 | 138 (69%) |
| None | 9 (5%) |
| Number of days of supplements prescribed |  |
| Median | 30 (28, 30) |
| 4 – 14 | 45 (22%) |
| 15 – 30 | 152 (76%) |
| 31 – 60 | 0 (0%) |
| 61 – 90 | 4 (2%) |
| Iron dose prescribed |  |
| Once daily | 139 (69%) |
| Twice daily | 60 (30%) |
| Twice weekly | 2 (1%) |
| Primary reason for prescribing iron supplements |  |
| Patient or caretaker requests it | 1 (0.5%) |
| Clinical condition warrants it | 106 (53%) |
| Lab test results warrant it | 92 (46%) |
| Other | 2 (1%) |
| Clinical features that influence iron supplement use in non-pregnant adults with HIV |  |
| Recent weight loss | 32 (16%) |
| Pallor | 168 (84%) |
| Recent hair loss | 38 (19%) |
| Nail deformities | 60 (30%) |
| Diarrhea | 36 (18%) |
| All adult HIV patients should receive supplements | 18 (9%) |
| None of the above | 9 (5%) |
| Clinical features that influence iron supplement use in pregnant adults with HIV |  |
| Recent weight loss | 11 (6%) |
| Pallor | 81 (43%) |
| Recent hair loss | 4 (2%) |
| Nail deformities | 17 (9%) |
| Diarrhea | 5 (3%) |
| All pregnant HIV patients should receive supplements | 154 (82%) |
| None of the above | 4 (2%) |
| Clinical features that influence iron supplement use in TB coinfection |  |
| Recent weight loss | 50 (25%) |
| Pallor | 158 (82%) |
| Recent hair loss | 36 (19%) |
| Nail deformities | 59 (31%) |
| Diarrhea | 27 (14%) |
| All patients coinfectd with TB patients should receive supplements | 36 (19%) |
| None of the above | 6 (3%) |
| Clinical features that influence iron supplement use in children |  |
| Recent weight loss | 53 (27%) |
| Pallor | 160 (87%) |
| Recent hair loss | 41 (22%) |
| Nail deformities | 48 (26%) |
| Diarrhea | 38 (21%) |
| All pediatric patients should receive supplements | 33 (18%) |
| None of the above | 0 (0%) |
| Clinician considers hemoglobin test to prescribe or discontinue iron supplements |  |
| Yes | 197 (98%) |
| Hemoglobin Cutoff below which iron supplements are initiated |  |
| <7 g/dl | 39 (20%) |
| <8.5 g/dl | 71 (36%) |
| <10 g/dl | 54 (27%) |
| <11 g/dl | 27 (14%) |
| Others | 7 (4%) |
| Lower cutoff below which iron supplements are discontinued and blood transfusion recommended instead |  |
| <7 g/dl | 41 (20%) |
| <8.5 g/dl | 26 (13%) |
| <10 g/dl | 26 (13%) |
| <11 g/dl | 49 (24%) |
| Others | 59 (29%) |
| Higher cutoff above which iron supplements discontinued |  |
| Yes | 168 (85%) |

The 193 clinicians who reported usual MMS prescription **(Table 3)** usually prescribed MMS supplements once daily (55%) for a median of 14 days (IQR: 14, 30). Patients’ clinical condition (84%) was the primary reason for prescribing the supplements. In addition to pallor (50%), unlike for iron supplements, clinicians considered other clinical signs more frequently when deciding whether to prescribe MMS to non-pregnant adults: recent hair loss (42%), recent weight loss (31%), diarrhea (28%) and nail deformities (25%). They considered these same signs more frequently for patients who were pregnant, co-infected with TB or were children. 100% of respondents believed that all pregnant women should receive MMS.

**Table 3.** Prescribing pattern of multivitamin supplements (N=193)

| Variable | N (%) |
| --- | --- |
| Number of patients prescribed MMS supplements in the last week |  |
| More than 20 | 18 (9%) |
| 10 to 20 | 31 (16%) |
| 5 to 9 | 120 (62%) |
| None | 25 (13%) |
| Number of days of supplements prescribed |  |
| Median | 14 (14, 30) |
| 3 – 14 | 97 (50%) |
| 15 – 30 | 92 (48%) |
| 31 – 60 | 0 (0%) |
| 61 – 90 | 4 (2%) |
| MMS dose prescribed |  |
| Once daily | 104 (55%) |
| Twice daily | 85 (45%) |
| Twice weekly | 2 (1%) |
| Primary reason for prescribing MMS supplements |  |
| Patient or caretaker requests it | 1 (1%) |
| Clinical condition warrants it | 162 (84%) |
| Lab test results warrant it | 30 (16%) |
| Clinical features that influence MMS supplement use in non-pregnant adults with HIV |  |
| Recent weight loss | 50 (31%) |
| Pallor | 81 (50%) |
| Recent hair loss | 69 (42%) |
| Nail deformities | 41 (25%) |
| Diarrhea | 45 (28%) |
| All adult HIV patients should receive supplements | 32 (20%) |
| None of the above | 17 (9%) |
| Clinical features that influence MMS supplement use in pregnant adults with HIV |  |
| Recent weight loss | 44 (23%) |
| Pallor | 58 (64%) |
| Recent hair loss | 43 (47%) |
| Nail deformities | 32 (35%) |
| Diarrhea | 33 (36%) |
| All pregnant HIV patients should receive supplements | 91 (100%) |
| None of the above | 15 (8%) |
| Clinical features that influence MMS supplement use in TB coinfection |  |
| Recent weight loss | 65 (34%) |
| Pallor | 85 (77%) |
| Recent hair loss | 54 (49%) |
| Nail deformities | 41 (37%) |
| Diarrhea | 40 (36%) |
| All patients coinfectd with TB patients should receive supplements | 111 (100%) |
| None of the above | 11 (6%) |
| Clinical features that influence MMS supplement use in children |  |
| Recent weight loss | 69 (37%) |
| Pallor | 81 (59%) |
| Recent hair loss | 47 (34%) |
| Nail deformities | 30 (22%) |
| Diarrhea | 39 (28%) |
| All patients coinfectd with TB patients should receive supplements | 49 (36%) |
| None of the above | 0 (0%) |
| Clinician considers hemoglobin test to prescribe or discontinue MMS supplements |  |
| Yes | 129 (68%) |
| Hemoglobin Cutoff below which MMS supplements are initiated |  |
| <7 g/dl | 18 (10%) |
| <8.5 g/dl | 72 (41%) |
| <10 g/dl | 37 (21%) |
| <11 g/dl | 13 (7%) |
| Others | 35 (20%) |
| Lower cutoff below which MMS supplements are discontinued and blood transfusion recommended instead |  |
| <7 g/dl | 95 (55%) |
| <8.5 g/dl | 14 (8%) |
| <10 g/dl | 2 (1%) |
| <11 g/dl | 15 (9%) |
| Others | 48 (28%) |
| Higher cutoff above which MMS supplements discontinued |  |
| Yes | 163 (85%) |

The 82 clinicians who reported usual RUTF prescription **(Table 4)** usually prescribed them as twice daily dose (61%) for a median of 17 days (IQR: 4, 70). Patients’ clinical condition (85%) and laboratory test results (13%) were the primary reasons for prescribing the supplements. Recent weight loss was the most commonly considered clinical feature among non-pregnant adults (84%), pregnant adults (75%), children (100%) and individuals co-infected with TB (100%), though other clinical signs and symptoms were considered too.

**Table 4.** Prescribing pattern of RUTF supplements (N=82)

| Variable | N (%) |
| --- | --- |
| Number of patients prescribed RUTF supplements in the last week |  |
| More than 20 | 5 (6%) |
| 10 to 20 | 8 (10%) |
| 5 to 9 | 56 (68%) |
| None | 13 (16%) |
| Number of days of RUTF prescribed |  |
| Median | 17 (4, 70) |
| 3 – 14 | 41 (50%) |
| 15 – 30 | 39 (48%) |
| 31 – 60 | 0 (0%) |
| 61 – 90 | 2 (2%) |
| RUTF dose prescribed |  |
| Once daily | 32 (39%) |
| Twice daily | 50 (61%) |
| Primary reason for prescribing RUTF supplements |  |
| Patient or caretaker requests it | 1 (1%) |
| Clinical condition warrants it | 70 (85%) |
| Lab test results warrant it | 11 (13%) |
| Clinical features that influence RUTF supplement use in non-pregnant adults with HIV |  |
| Recent weight loss | 33 (65%) |
| Pallor | 28 (55%) |
| Recent hair loss | 32 (63%) |
| Nail deformities | 11 (22%) |
| Diarrhea | 16 (31%) |
| All adult HIV patients should receive supplements | 0 (0%) |
| None of the above | 7 (8%) |
| Clinical features that influence RUTF supplement use in pregnant adults with HIV |  |
| Recent weight loss | 63 (75%) |
| Pallor | 35 (60%) |
| Recent hair loss | 33 (57%) |
| Nail deformities | 10 (17%) |
| Diarrhea | 16 (28%) |
| All pregnant HIV patients should receive RUTF | 0 (0%) |
| None of the above | 7 (8%) |
| Clinical features that influence RUTF supplement use in TB coinfection |  |
| Recent weight loss | 61 (100%) |
| Pallor | 34 (62%) |
| Recent hair loss | 37 (67%) |
| Nail deformities | 10 (18%) |
| Diarrhea | 22 (40%) |
| All patients coinfectd with TB patients should receive supplements | 0 (0%) |
| None of the above | 1 (1%) |
| Clinical features that influence RUTF supplement use in children |  |
| Recent weight loss | 56 (100%) |
| Pallor | 28 (56%) |
| Recent hair loss | 33 (66%) |
| Nail deformities | 8 (16%) |
| Diarrhea | 23 (46%) |
| All patients coinfectd with TB patients should receive supplements | 0 (0%) |
| None of the above | 0 (0%) |
| Clinician considers physical exam findings to prescribe or discontinue RUTF supplements |  |
| Yes | 77 (96%) |
| Physical exam findings that guide initiation of RUTF |  |
| BMI too low | 46 (56%) |
| WHZ scores suggest patient is wasted | 20 (24%) |
| Weight too small | 16 (20%) |

For all three types of supplements considered, clinicians were most concerned (approximately 20% or greater) about the need for frequent refills and of supplements being out-of-stock **(Figure 2)**. Some clinicians were concerned about the cost of the supplements. Infection risk (3%) and contraindications (7%) were also concerns that a small number of clinicians had that influenced their prescribing patterns for iron supplements.

**Figure 2.**
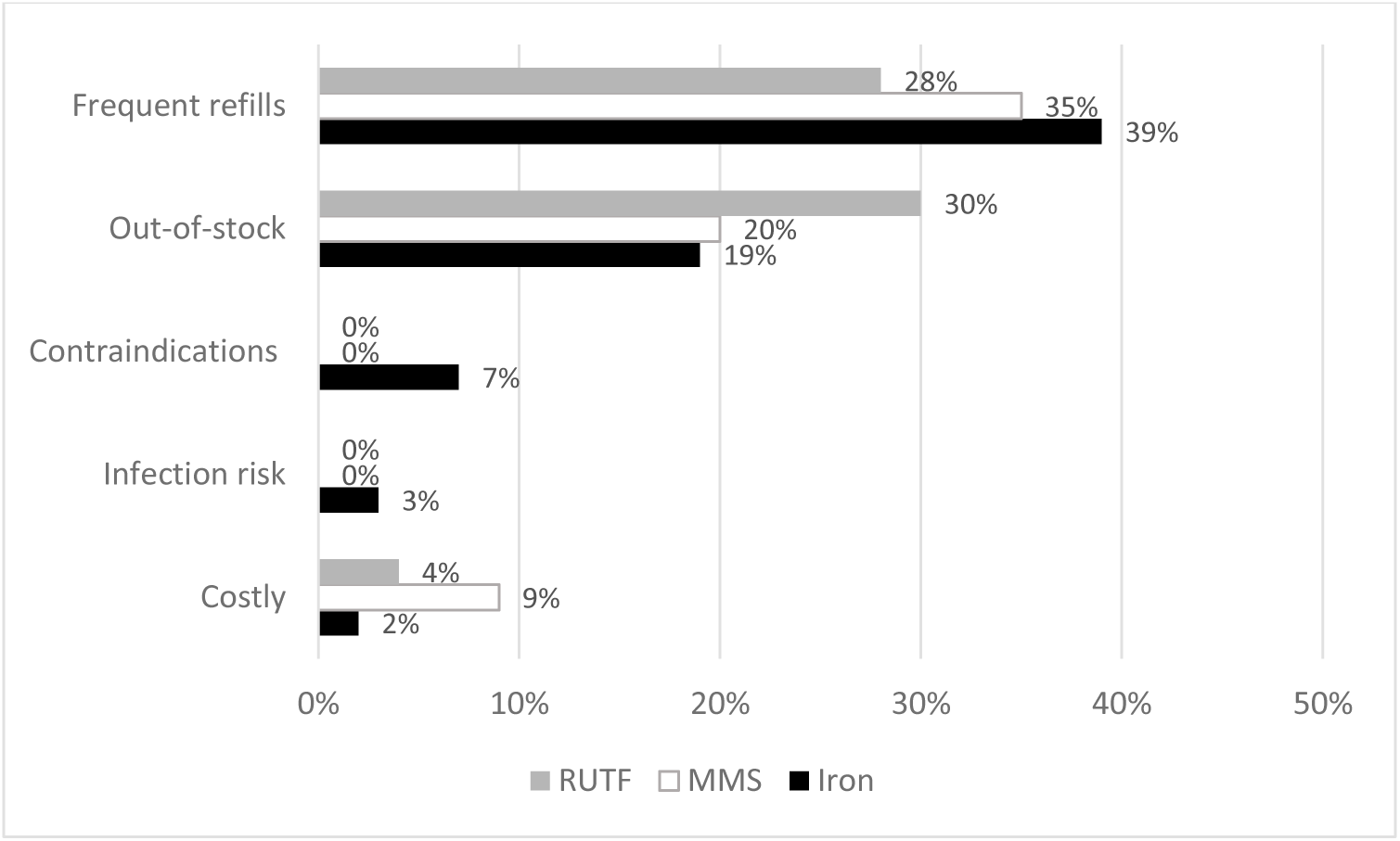
Barriers to prescribing nutritional supplements.

Univariable models evaluating the relationships of multiple demographic and professional characteristics with clinicians’ prescription of nutritional interventions found that the clinicians who self-reported attended to between 10 and <100 patients in the preceding week were more likely to prescribe RUTF, compared to clinicians who attended to <10 patients weekly**(Supplementary table 1)**.

No other significant relationships for iron, MMS or RUTF with demographic and professional factors was found. Factors that were evaluated include the district, facility type, clinicians’ age, qualification, number of years since graduation, years since working at MDH, the number of patients seen in the last week.

For each of non-pregnant adult, pregnant adult, TB-HIV coinfected and pediatric populations, univariable models evaluated the relationship of self-reported reasons for prescribing supplements and clinicians’ prescription of nutritional interventions. Clinicians who believed that all pregnant HIV patients should receive iron supplements were 1.23-fold (95% CI: 1.03, 1.56) more likely to report prescribing them**(Supplementary table 2)**. No other significant relationships were identified.

## Discussion

We conducted a survey of clinicians’ prescription of iron supplements, MMS and RUTF at HIV care and treatment centers in Dar es Salaam, and found that these nutritional interventions were frequently recommended by clinicians to their patients. We identified the common patterns of prescribing the different supplements, including the reasons for prescribing, duration and dose. Most clinicians believed that pregnant HIV patients should receive iron supplements and MMS. All clinicians believed that all patients coinfected with TB and HIV should receive MMS. Only 3% considered the presence of signs of infections to be contraindications to iron supplements. Instead, clinicians were concerned that stockouts and the need for frequent refills discourage them from prescribing iron supplements.

The overwhelming majority of clinicians in this network of HIV clinics reported that they usually prescribe iron supplements to PLHIV. There are no current WHO guidelines for iron supplementation among PLHIV due to concerns that iron supplementation may be unsafe or ineffective in this context^(17)^. These concerns are based on the understanding that both iron deficiency and iron overload may be associated with worsening immune function^(28)^. Epidemiological studies, including studies set in the same network of clinics as our survey, have shown increased risk of mortality when iron status is elevated ^(18; 19; 20)^. These effects possibly result from faster HIV replication, more frequent opportunistic infections, and oxidative damage to tissues due to elevated iron status and higher levels of non-transferrin bound iron^(29; 30; 31; 32)^. As shown in our study, clinicians self-report that they prescribe iron supplements to PLHIV, possibly due to their concerns about iron deficiency. Iron deficiency is highly prevalent (7% – 48%) among PLHIV^(18; 20; 33; 34)^. Severe iron deficiency leads to anemia, which also increases mortality risk. Iron supplementation is well-known to be effective in the general population to prevent and treat anemia^(35; 36; 37)^. Iron supplementation has however been poorly studied among PLHIV^(21; 22; 23; 24)^. Clinicians who didn’t prescribe iron supplements were mostly concerned about stock-outs and frequent need for refills. Only few clinicians were concerned about the infection-related risks. Nonetheless, well-designed studies are required to evaluate the safety and effectiveness of iron supplementation among adult PLHIV to guide clinical and public health decision making.

Most clinicians in our study also reported they frequently prescribed MMS. Only 13% said they did not prescribe the previous week. Randomized trials and observational studies among ART-naïve and ART-experienced individuals have demonstrated that MMS reduces the risk of mortality, HIV disease progression, incident tuberculosis and immunologic failure^(13; 38; 39; 40)^. Micronutrient deficiencies are very common among PLHIV; often with multiple concurrent micronutrient deficiencies^(41; 42)^. These deficiencies result from increased food security risk, reduced dietary intake due to anorexia, decreased nutrient absorption from systemic inflammation, altered metabolism, and nutrient losses from diarrhea^(42)^. Clinicians in our study were more likely to prescribe MMS for therapeutic purposes and for short durations, to address the effects of micronutrient deficiencies, rather than for prophylactic reasons. Only 20% of the clinicians believed that MMS should be prescribed to all PLHIV. Clear evidence-based guidelines regarding the use of MMS in HIV could potentially improve their appropriate use among PLHIV.

RUTF is effective to treat acute malnutrition among PLHIV on ART^(43)^. Clinicians in our study prescribed RUTF often, and 34% prescribed RUTF in the preceding week. The prescriptions were often for guided by patients’ clinical condition, especially in the presence of recent weight loss or hair loss, and for durations lasting 30 days or less. No clinician recommended universal use. Taken together, there seemed to be great alignment among clinicians regarding whether and when to prescribe RUTF to their patients. There was less alignment with iron and multiple micronutrient supplementation. The lack of global recommendations or guidelines regarding the use of micronutrient supplements likely explains the disparity. Possible alternative explanations may be that the clinicians perceive the effectiveness of micronutrient supplements to be limited, relative to macronutrient RUTF supplementation.

Clinicians were concerned about patient access for the supplements they intended to prescribe. They were concerned about stockouts for all the supplements. Stockouts are a common hindrance to the uptake and regular use of micronutrient supplementation in developing countries^(44; 45; 46)^. Addressing these supply chain challenges could potentially improve patients’ quality of life, and reduce the risk of outcomes. Private facilities could play a key role to fill the shortfall from the HIV care and treatment program^(47)^. Clinicians were also worried about the need for frequent refills, a result of the transient effect of these supplements and the supply chain issues mentioned above. Only few clinicians were concerned about the cost of the supplements, as iron and multivitamins tend to be cheap, and RUTF has been provided free of charge to patients.

Our study has one important limitation. Participants were asked to self-report their prescription patterns of micronutrient supplementation. Clinicians sometimes prescribed iron and multiple micronutrient supplements without recording in the patients’ health records, especially when the patients have to purchase the supplements at private pharmacies.

Although RUTFs were always documented in the health records, the anonymous nature of the survey makes it impossible to validate the prescribing patterns. It was also impossible to confirm the clinical and laboratory indications for which supplements would have been prescribed.

## Conclusion

We conducted a cross-sectional survey among clinicians with prescribing responsibility at clinics providing care to PLHIV in Dar es-Salaam, Tanzania. We found that substantial proportion prescribe nutritional supplements, although there is a lot of variation in why they prescribe them, and for how long. Clear guidelines regarding the prescription of nutritional supplements may be necessary given the state of the evidence regarding the safety and effectiveness of the supplements.

## Data Availability

All data produced in the present study are available upon reasonable request to the author

## Abbreviations used in text

AIDS: Acquired Immune Deficiency Syndrome
ART: Antiretroviral therapy
BMI: Body Mass Index
CI: Confidence Interval
HAART: Highly Active Antiretroviral Therapy
HIV: Human Immunodeficiency Virus
ID: Iron deficiency
IDA: Iron deficiency anemia
IQR: Interquartile range
PLHIV: People living with HIV
RR: Relative risk
TB: Tuberculosis;

---

**Supplementary Table 1.** Demographic and professional characteristics associated with clinician’s prescription of nutritional interventions.

|  | Iron |  |  | MMS |  |  | RUTF |  |  |
| --- | --- | --- | --- | --- | --- | --- | --- | --- | --- |
|  | Yes,<br>n (%) | No,<br>n (%) | Prev Ratio (95% CI) | Yes,<br>n (%) | No,<br>n (%) | Prev Ratio (95% CI) | Yes,<br>n (%) | No,<br>n (%) | Prev Ratio (95% CI) |
| District |  |  |  |  |  |  |  |  |  |
| Ilala | 10 (15%) | 56 (29%) | Ref | 13 (14%) | 53 (32%) | Ref | 2 (11%) | 64 (27%) | Ref |
| Kigamboni | 2 (29%) | 5 (3%) | 1.01 (0.39, 2.16) | 1 (1%) | 6 (4%) | 0.69 (0.21, 1.67) | 1 (6%) | 6 (3%) | 4.71 (0.24, 43.9) |
| Kinondoni | 28 (35%) | 52 (27%) | 0.88 (0.61, 1.27) | 33 (37%) | 47 (29%) | 0.95 (0.66, 1.36) | 7 (39%) | 73 (31%) | 2.89 (0.73, 18.9) |
| Temeke | 21 (31%) | 46 (24%) | 1.04 (0.72, 1.50) | 21 (23%) | 46 (28%) | 0.82 (0.55, 1.22) | 4 (22%) | 63 (27%) | 1.97 (0.40, 13.9) |
| Ubungo | 1 (3%) | 33 (17%) | 0.73 (0.43, 1.18) | 22 (24%) | 12 (7%) | 0.92 (0.57, 1.45) | 4 (22%) | 30 (13%) | 3.88 (0.80, 27.0) |
| Facility type |  |  |  |  |  |  |  |  |  |
| ANC | 3 (5%) | 1 (11%) | 1.28 (0.39, 3.03) | 2 (2%) | 2 (1%) | 0.68 (0.11, 2.14) | 0 (0%) | 4 (2%) | NR |
| CTC | 37 (60%) | 125 (65%) | Ref | 52 (58%) | 110 (67%) | Ref | 13 (72%) | 149 (63%) | Ref |
| TB/HIV clinic | 8 (13%) | 13 (7%) | 1.15 (0.69, 1.82) | 8 (9%) | 13 (8%) | 1.12 (0.81, 1.53) | 2 (11%) | 65 (28%) | 1.78 (0.43, 5.00) |
| Other | 14 (23%) | 53 (28%) | 0.99 (0.71, 1.36) | 28 (31%) | 39 (24%) | 1.17 (0.69, 1.86) | 3 (17%) | 18 (8%) | 0.37 (0.06, 1.30) |
| Age |  |  |  |  |  |  |  |  |  |
| <30 | 7 (12%) | 25 (14%) | Ref | 12 (14%) | 20 (13%) | Ref | 2 (13%) | 30 (14%) | Ref |
| 30 – <40 | 12 (21%) | 42 (23%) | 1.06 (0.84, 1.40) | 23 (26%) | 31 (21%) | 0.93 (0.58, 1.51) | 4 (25%) | 50 (22%) | 1.23 (0.17, 7.07) |
| 40 – <50 | 25 (44%) | 69 (38%) | 1.06 (0.87, 1.39) | 38 (43%) | 56 (37%) | 0.95 (0.62, 1.48) | 7 (44%) | 87 (8%) | 1.46 (0.33, 7.12) |
| >50 | 13 (23%) | 46 (25%) | 1.04 (0.83, 1.37) | 15 (17%) | 44 (29%) | 0.70 (0.43, 1.15) | 3 (18%) | 56 (25%) | 1.46 (0.42, 6.59) |
| Qualification |  |  |  |  |  |  |  |  |  |
| Clinical officer | 39 (63%) | 127 (66%) | Ref | 60 (67%) | 106 (65%) | Ref | 12 (67%) | 154 (65%) | Ref |
| Medical assistant | 19 (31%) | 35 (18%) | 1.11 (0.79, 1.54) | 18 (20%) | 36 (22%) | 0.88 (0.61, 1.26) | 3 (17%) | 51 (22%) | 0.82 (0.28, 3.46) |
| Medical doctor | 4 (7%) | 30 (16%) | 0.94 (0.60, 1.41) | 12 (13%) | 22 (13%) | 0.89 (0.56, 1.34) | 3 (17%) | 31 (13%) | 0.63 (0.12, 3.24) |
| Years since graduation |  |  |  |  |  |  |  |  |  |
| 1 – 9 | 27 (46%) | 84 (44%) | Ref | 43 (49%) | 68 (42%) | Ref | 8 (44%) | 103 (44%) | 0.97 (0.34, 3.08) |
| 10 – 19 | 16 (27%) | 51 (27%) | 1.08 (0.77, 1.51) | 24 (27%) | 43 (26%) | 0.95 (0.67, 1.34) | 5 (28%) | 62 (27%) | Ref |
| 20 – 29 | 14 (24%) | 44 (23%) | 0.98 (0.68, 1.40) | 16 (18%) | 42 (26%) | 0.92 (0.63, 1.33) | 5 (28%) | 53 (23%) | 1.16 (0.34, 3.97) |
| 30 – 39 | 2 (3%) | 11 (6%) | 1.09 (0.55, 1.96) | 5 (6%) | 8 (5%) | 0.98 (0.48, 1.800) | 0 (0%) | 13 (6%) | NR |
| 40 – 50 | 0 (0%) | 2 (1%) | 1.29 (0.21, 4.08) | 0 (0%) | 2 (1%) | 1.28 (0.21, 4.03) | 0 (0%) | 2 (1%) | NR |
| Years since working in HIV care |  |  |  |  |  |  |  |  |  |
| 0 – 5 | 30 (48%) | 102 (53%) | Ref | 51 (57%) | 81 (49%) | Ref | 10 (56%) | 122 (52%) | Ref |
| 6 – 10 | 14 (23%) | 42 (22%) | 1.22 (0.86, 1.70) | 19 (21%) | 37 (23%) | 0.91 (0.64, 1.28) | 5 (28%) | 61 (26%) | 0.71 (0.16, 2.21) |
| 11 – 16 | 18 (29%) | 48 (25%) | 1.13 (0.81, 1.57) | 20 (22%) | 46 (28%) | 1.10 (0.77, 1.54) | 3 (17%) | 53 (23%) | 1.00 (0.32, 2.69) |
| Number of patients seen preceding week |  |  |  |  |  |  |  |  |  |
| <10 | 10 (17%) | 34 (18%) | Ref | 20 (23%) | 24 (15%) | Ref | 2 (12%) | 42 (18%) | Ref |
| 10 – <30 | 21 (35%) | 55 (29%) | 1.20 (0.78, 1.87) | 26 (29%) | 50 (31%) | 1.04 (0.68, 1.60) | 1 (6%) | 75 (33%) | 1.95 (1.03, 4.26) |
| 30 – <60 | 4 (7%) | 30 (16%) | 1.12 (0.66, 1.90) | 11 (12%) | 23 (15%) | 0.98 (0.58, 1.64) | 5 (29%) | 29 (13%) | 2.10 (1.01, 4.78) |
| 60 – <100 | 3 (5%) | 24 (13%) | 1.20 (0.68, 2.06) | 11 (12%) | 16 (10%) | 1.19 (0.69, 2.00) | 3 (18%) | 24 (10%) | 2.24 (1.04, 5.14) |
| >100 | 22 (37%) | 45 (24%) | 1.27 (0.82, 2.00) | 21 (24%) | 46 (29%) | 1.00 (0.64, 1.56) | 6 (35%) | 61 (26%) | 1.83 (0.94, 4.05) |

**Supplementary Table 2.** Self-reported reasons for prescribing and their relationship with clinician’s prescription of nutritional interventions.

|  | Iron |  |  | MMS |  |  | RUTF |  |  |
| --- | --- | --- | --- | --- | --- | --- | --- | --- | --- |
|  | Yes,<br>n (%) | No,<br>n (%) | Prev Ratio (95% CI) | Yes,<br>n (%) | No,<br>n (%) | Prev Ratio (95% CI) | Yes,<br>n (%) | No,<br>n (%) | Prev Ratio (95%<br>CI) |
| Clinical features to that influence supplement use in non-pregnant adults with HIV |  |  |  |  |  |  |  |  |  |
| Recent weight loss | 32 (16%) | 10 (19%) | 0.95 (0.77, 1.12) | 31 (16%) | 11 (18%) | 0.96 (0.77, 1.14) | 14 (17%) | 28 (17%) | 1.02 (0.60, 1.57) |
| Pallor | 168 (84%) | 42 (81%) | 1.06 (0.90, 1.32) | 16 (84%) | 50 (83%) | 1.01 (0.85, 1.26) | 71 (88%) | 135 (81%) | 1.41 (0.85, 2.70) |
| Recent hair loss | 38 (19%) | 12 (23%) | 0.95 (0.78, 1.10) | 37 (19%) | 13 (22%) | 0.96 (0.78, 1.14) | 17 (21%) | 32 (19%) | 1.07 (0.67, 1.60) |
| Nail deformities | 60 (30%) | 12 (23%) | 1.07 (0.93, 1.21) | 56 (29%) | 16 (27%) | 1.03 (0.88, 1.19) | 20 (25%) | 51 (31%) | 0.81 (0.52, 1.21) |
| Diarrhea | 36 (18%) | 14 (27%) | 0.89 (0.72, 1.04) | 34 (18%) | 16 (27%) | 0.87 (0.69, 1.04) | 18 (22%) | 30 (18%) | 1.18 (0.75, 1.75) |
| All patients | 18 (9%) | 2 (4%) | NR | 17 (9%) | 3 (5%) | 1.13 (0.86, 1.32) | 7 (9%) | 13 (8%) | 1.07 (0.51, 1.82) |
| Clinical features that influence iron supplement use in pregnant adults with HIV |  |  |  |  |  |  |  |  |  |
| Recent weight loss | 11 (6%) | 4 (8%) | 0.92 (0.60, 1.15) | 13 (7%) | 2 (3%) | NR | 4 (5%) | 10 (6%) | 0.86 (0.29, 1.70) |
| Pallor | 81 (43%) | 19 (40%) | 1.03 (0.90, 1.17) | 72 (40%) | 28 (49%) | 0.92 (0.78, 1.06) | 36 (46%) | 63 (41%) | 1.12 (0.78, 1.60) |
| Recent hair loss | 4 (2%) | 3 (6%) | 0.71 (0.28, 1.09) | 5 (3%) | 2 (4%) | 0.94 (0.46, 1.26) | 2 (3%) | 5 (3%) | 0.83 (0.16, 1.95) |
| Nail deformities | 17 (9%) | 5 (10%) | 0.97 (0.71, 1.17) | 17 (10%) | 5 (9%) | 1.02 (0.75, 1.24) | 9 (11%) | 12 (8%) | 1.29 (0.69, 2.03) |
| Diarrhea | 5 (3%) | 3 (6%) | 0.78 (0.45, 1.34) | 5 (3%) | 3 (5%) | 0.82 (0.48, 1.41) | 3 (4%) | 5 (3%) | 1.10 (0.32, 2.16) |
| All patients | 154 (82%) | 32 (67%) | 1.23 (1.03, 1.56) | 140 (79%) | 46 (81%) | 0.97 (0.83, 1.18) | 62 (79%) | 122 (80%) | 0.93 (0.63, 1.49) |
| Clinical features that influence iron supplement use in TB coinfection |  |  |  |  |  |  |  |  |  |
| Recent weight loss | 50 (25%) | 8 (15%) | NR | 44 (23%) | 14 (23%) | 0.99 (0.82, 1.15) | 18 (22%) | 38 (23%) | 0.97 (0.61, 1.45) |
| Pallor | 158 (82%) | 39 (81%) | 1.01 (0.88, 1.24) | 145 (80%) | 52 (88%) | 0.88 (0.76, 1.05) | 62 (81%) | 131 (82%) | 0.92 (0.61, 1.53) |
| Recent hair loss | 36 (19%) | 6 (13%) | NR | 32 (18%) | 10 (17%) | 1.01 (0.81, 1.19) | 16 (21%) | 25 (16%) | 1.25 (0.77, 1.86) |
| Nail deformities | 59 (31%) | 7 (15%) | NR | 52 (29%) | 14 (24%) | 1.06 (0.90, 1.23) | 23 (30%) | 41 (26%) | 1.14 (0.75, 1.67) |
| Diarrhea | 27 (14%) | 6 (13%) | 1.03 (0.83, 1.19) | 22 (12%) | 11 (19%) | 0.97 (0.64, 1.07) | 13 (17%) | 19 (12%) | 1.29 (0.76, 1.97) |
| All patients | 36 (19%) | 8 (17%) | 1.03 (0.85, 1.18) | 37 (20%) | 7 (12%) | 1.14 (0.96, 1.32) | 15 (20%) | 29 (18%) | 1.06 (0.64, 1.61) |
| Clinical features that influence iron supplement use in children |  |  |  |  |  |  |  |  |  |
| Recent weight loss | 53 (27%) | 7 (14%) | NR | 48 (26%) | 12 (21%) | 1.06 (0.90, 1.23) | 21 (26%) | 37 (23%) | 1.12 (0.73, 1.64) |
| Pallor | 160 (87%) | 44 (94%) | NR | 152 (86%) | 52 (95%) | NR | 68 (88%) | 132 (87%) | 1.06 (0.64, 2.05) |
| Recent hair loss | 41 (22%) | 9 (19%) | 1.04 (0.87, 1.19) | 35 (20%) | 15 (27%) | 0.90 (0.72, 1.07) | 17 (22%) | 31 (21%) | 1.06 (0.66, 1.59) |
| Nail deformities | 48 (26%) | 7 (15%) | NR | 43 (24%) | 12 (22%) | 1.03 (0.86, 1.20) | 19 (25%) | 34 (23%) | 1.08 (0.69, 1.60) |
| Diarrhea | 38 (21%) | 9 (19%) | 1.02 (0.85, 1.17) | 32 (18%) | 15 (27%) | 0.87 (0.68, 1.05) | 20 (26%) | 26 (17%) | 1.39 (0.91, 2.01) |
| All pediatric patients | 33 (18%) | 5 (11%) | NR | 34 (19%) | 4 (7%) | NR | 13 (17%) | 25 (17%) | 1.02 (0.59, 1.57) |
| Provides nutritional counseling |  |  |  |  |  |  |  |  |  |
| Yes | 3 (6%) | 30 (15%) | NR | 26 (14%) | 7 (12%) | 1.04 (0.82, 1.22) | 11 (13%) | 21 (13%) | 1.02 (0.59, 1.57) |
| No | 49 (94%) | 171 (85%) | Ref | 167 (87%) | 53 (88%) | Ref | 71 (87%) | 146 (87%) | Ref |

